# Can we use these masks? Rapid Assessment of the Inhalation Resistance Performance of Uncertified Medical Face Masks in the Context of Restricted Resources Imposed during a Public Health Emergency

**DOI:** 10.1101/2020.05.13.20100602

**Authors:** Steven Begg, Nwabueze Emekwuru, Nicolas Miché, Bill Whitney, Obuks Ejohwomu

## Abstract

In the case of a public health emergency such as the COVID-19 pandemic, access to large quantities of appropriate personal protection equipment (PPE) has presented a significant problem. A shortage of face masks and respirators has been widely reported across the world. A concerted effort to manufacture high volumes has not unsurprisingly put pressure on the supply chain and the important certification processes. PPE procured or donated as uncertified stock requires rigorous, expedient and scientifically informed evidence before decisions can be made regarding suitable deployment, expensive certification, return or possible destruction of stock. This paper reports a series of experiments devised in reaction to this situation. In this study, an experimental methodology for the assessment of the filtration performance of samples of real-world, uncertified, fluid resistant surgical masks (FRSM type IIR) was evaluated in the resource limited (lockdown) environment of the COVID-19 pandemic. A steady-state flow rig was adapted to incorporate a bespoke filter flow chamber for mounting face masks in order to evaluate the resistance to air flow as an indicator of mask inhalation performance. Pure air was drawn through a specified control surface area at known flow rate conditions; the resistance to the air flow through the masks was measured as the resulting pressure drop. Over 60 tests were performed from 4 different, randomly sampled batches and compared to a control sample of EN Type IIR certified FRSM masks. Steady-state volumetric airflow rates of 30 and 95 lmin^−1^ were chosen to represent deep breathing and vigorous exercise conditions respectively. The results showed that the sample masks produced a pressure drop of between 26% to 58% compared to the control batch at the lower flow rate and 22% to 55% at the higher rate. The results for each sample were consistent across both flow rates. Within the group of masks tested, two sets (between 48% and 58% of the reference set) showed the potential to be professionally assessed for appropriate deployment in a suitable setting. Although the absolute values of pressure drop measured by this method are unlikely to correlate with other testing approaches, the observed, indicative trends and relative performance of the masks is key to this approach. Critically, this method does not replace certification but it has enabled a public body to quickly make decisions; certify, re-assign, refund, thus saving time and resources. The total time spent conducting the tests was less than 8 hours and the low cost method proposed can be repurposed for low resource regions.

## Introduction

The COVID-19 pandemic has led to a substantial increase in demand of personal protection equipment (PPE), including face masks and respirators. A shortage in supplies has particularly affected health workers who are at most risk of infection (Soucheray, 2020). The short fall has led to a rapid increase in the production of face masks motivated by the need for public and health care authorities to acquire hitherto unprecedented numbers of masks, from varying sources, to meet the acute demand. However, operational restrictions placed upon the supply chain have meant that many of these face masks are without certification. Authorities can be left with vast quantities that have to be either used as uncertified (putting health workers at risk) or put through a lengthy certification process, thereby delaying the deployment of the masks and incurring additional certification expenses. In the worst cases, masks may need to be returned (and potentially not refunded) or destroyed, thus incurring costs and without addressing the shortage.

Several National and International certification protocols exist for masks and respirators (see Table 1).

**Table 1:**
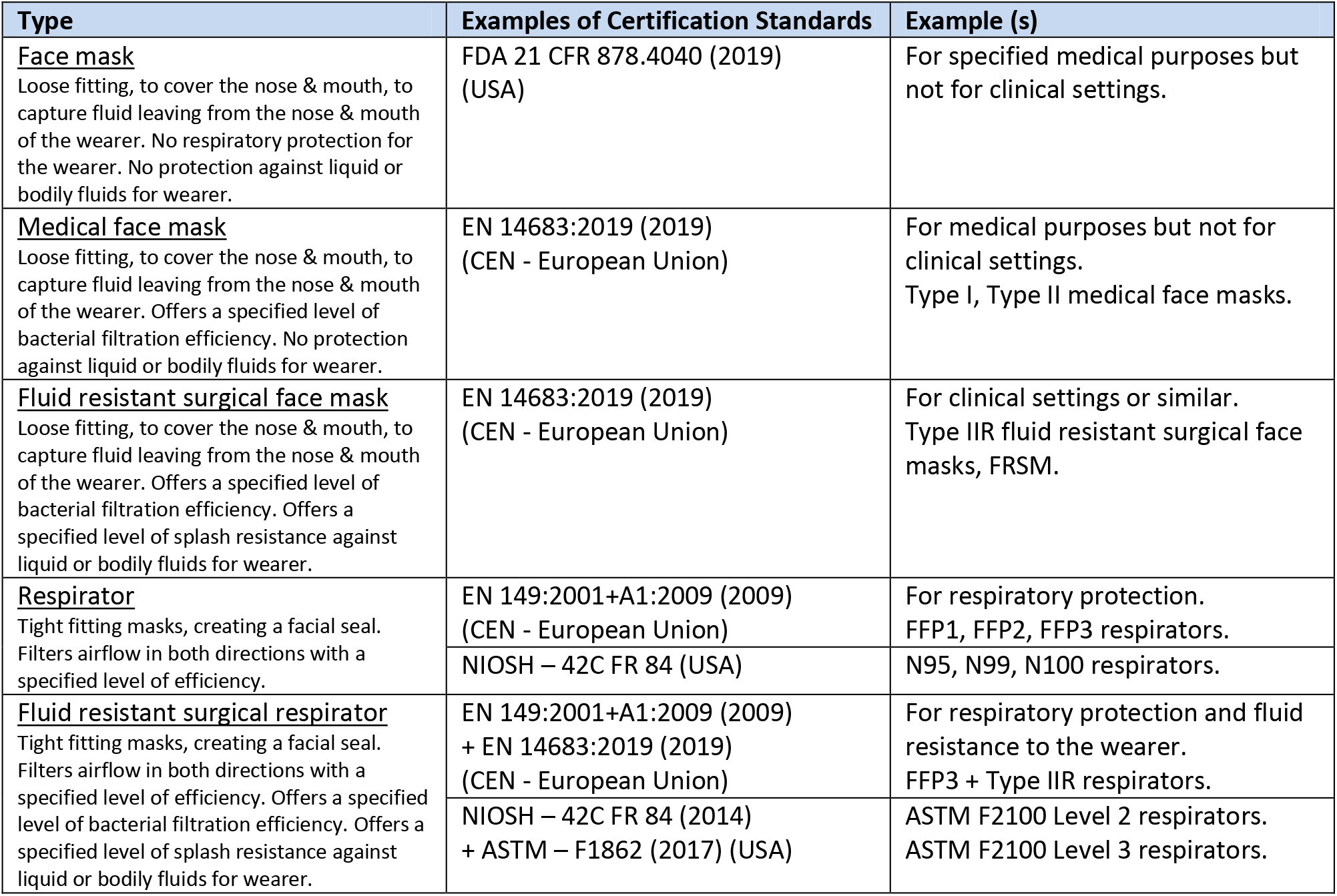
Examples of current masks and respirators and the relevant certification standards.

The processes involved in certification and approval can be lengthy and costly (CDC, 2018) either for the manufacturer, importer, institution or authority looking to distribute the devices. During a public health emergency, such as the current pandemic, limited available resources makes it increasingly difficult to carry out these procedures effectively. In such a scenario, access to appropriate, expedient and reliable scientific test data is an essential part of the process that leads to an informed decision; whether to send the devices for certification or to employ them in suitable settings that do not require certification or are considered of lower risk.

The shortage of masks that resulted as a consequence of previous epidemics has focused attention upon the testing of the filtration efficiency of home-made or uncertified masks and respirators made from different materials (Cooper et al. 1983; Rengasamy et al. 2010; Jung et al. 2013; Konda et al. 2020). Likewise, the re-use performance of the masks or respirators after some form of cleaning or sterilization (Viscusi et al. 2009; Liao et al. 2020) and alternative qualitative mask fit testing methods (Provenzano et al. 2020) have been studied. However, very little consideration has been given to devising rapid, scalable emergency protocols to help assess the performance of replacement, alternative masks or respirators, during the critical initial stages or beyond, in situations where emergency-use authorisation has been given due to the scarcity of supply. In a recent study, Mueller and Fernandez, (2020) propose a rapid screening tool to evaluate the filtration efficiency of masks when subjected to airborne particulates, similar in size to viruses. Approximately 2 hours of testing was required per mask, thereby limiting its suitability to an emergency rapid response type situation. This paper describes a method that was developed to rapidly assess the inhalation filtration performance of nose and mouth coverings (suitable for both medical face masks and respirators) in response to a public health emergency in a low resource and restricted (lockdown) scenario during the COVID-19 pandemic. It should be clear here that the tests were not devised as a means of certification; instead, a comparative measure of the performance of the masks against an approved Comité Européen de Normalization (CEN) EN certified benchmark. An important balance was sought between accuracy of results and test duration given the time constraints imposed by the pandemic emergency call.

## Experimental methodology

### Materials

The filtration performance of the medical masks was assessed by measuring the inhalation volumetric flow resistance to the passage of airflow under steady-state flow conditions. The resistive force was measured as a differential pressure drop across a specified surface area of the masks. The reproducibility and duration of the tests, given the extremely short operational time constraints, formed an important part of the study. The masks, apparatus and methodology are outlined in the following sections.

#### Masks

The FRSM type IIR masks used in this study were randomly sampled from large indeterminate or uncertified stock procured by a public body, from four different sources, during a lockdown week of the COVID-19 public health emergency. Ten masks were randomly selected from each individual batch and colour coded. From these, four were randomly selected for testing. The details of the masks are given in Table 2 along with approximate dimensions (for the range of small, medium and large sizes). A fifth sample of CEN EN certified Type IIR FRSM masks was selected in the same way. These were used as the control reference samples. All the uncertified masks were similar in appearance and composition to CEN approved EN Type IIR masks. They were made up of a conventional 3-ply (three layer) pleated design, but it was not possible by visual inspection to determine whether the middle layer was constructed of the standard “melt-blown” material. All of the masks included glued on ear loops (excepting the CEN EN approved masks which were stitched on) and sewn in wires to enable the wearer to adjust the fit to the bridge of the nose. The approximate sizes of the masks ranged from 167 to 180 mm in width and 90 to 95 mm in height.

**Table 2:** Details of the control and sample masks.

| Mask | Details | Number of samples selected from batch |
| --- | --- | --- |
| Control | EN 14683:2019 certified fluid resistant Type IIR surgical mask. Approximate size 180 x 90 mm (W x H) | 4 |
| A | Supplied surgical mask – no product information. With ear loops. 3-ply material. Approximate size 180 x 90 mm (W x H) | 4 |
| B | Supplied surgical mask - no product information. With ear loops. 3-ply material. Approximate size 167 x 90 mm (W x H) | 4 |
| C | Donated surgical mask - no product information. With ear loops. 3-ply material. Approximate size 167 x 95 mm (W x H) | 4 |
| D | Donated surgical mask - no product information. With ear loops. 3-ply material. Approximate size 167 x 90 mm (W x H) | 4 |

### Apparatus

A steady-state flow rig was adapted for the experimental evaluation of the filtration performance of the surgical masks. A schematic of the flow rig is shown in Figure 1 below.

**Figure 1:**
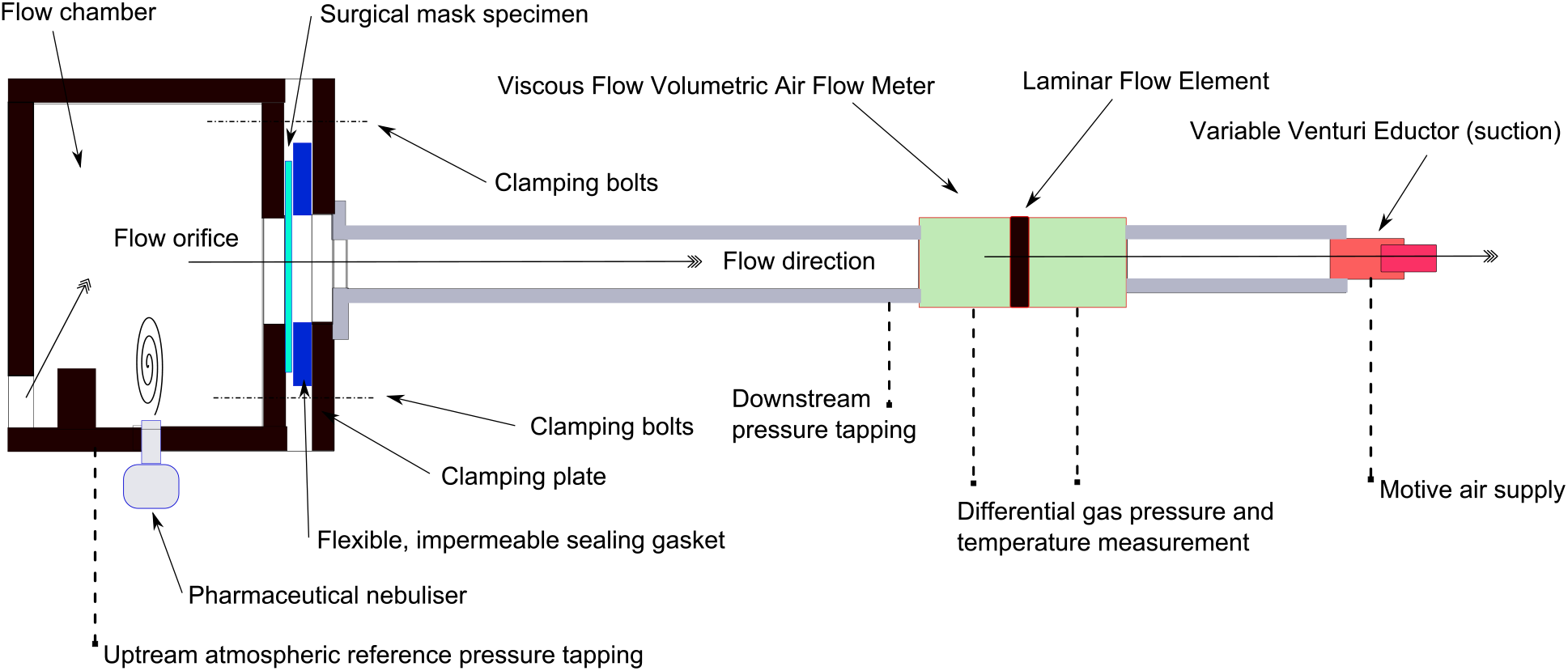
Schematic of Steady-State Flow Rig Adapted for PPE Mask Tests.

The 3 m test rig comprises a flow chamber for mounting the masks, a 1.5 m long developing flow pipe, a cylindrical volumetric air flow meter and a cylindrical flow rate regulator (eductor) to set the required flow rate. The variable venturi eductor was manufactured inhouse and is supplied with motive air from the laboratory air compressors. This was oil free, dry air, regulated to 4 bar for all the tests. The device can be adjusted on a fine thread to vary the venturi throat diameter and magnitude of vacuum created at the outlet. The low pressure at the exit of the test rig draws air through the flow chamber and viscous flow air meter (Alcock/Ricardo VFAM) in the direction indicated in the figure. An automotive throttle valve positioned downstream of the VFAM could be used to fine tune the flow rate. The main components are summarised as follows:

#### Flow chamber and mounting plates

The flow chamber was designed to allow the free flow of air into the test rig from the laboratory. Air entering through the bottom of the chamber is deflected upwards, in a forward tumbling motion, by a small step, designed to promote homogenous mixing in the chamber for tests incorporating water injection (a hole in the bottom of the chamber was provided for insertion of a nebuliser chamber). The mounting plates were designed to clamp the masks flat and accurately in place and facilitate exchanges. The plates were held tightly in place using four coach bolts. A reference flow orifice area was chosen for the tests as shown in Figure 2 for the FRSM Type IIR masks. This was cut into the mounting plates as a template. A firm, impermeable, foam gasket with a thickness of 1 cm was adapted to accommodate the shape of the hard nose ridge piece (within blue strip) in order to eliminate potential flow leakage. The mounting plate design incorporated plates for both flat (FRSM) and conical (N95) shaped masks/respirators. A series of screws on the side of the chamber were used to locate the ear loops. The pressure difference between the atmospheric, ambient conditions in the flow chamber and a pressure tapping downstream of the masks was recorded in mm of H_2_0 using a spirit levelled, liquid paraffin, inclined manometer with a scaling factor of 0.2. (Airflow Developments Limited, type 504). The accuracy in estimating the liquid level was +/− 1mm of H_2_0, equivalent to +/− 2 Pa on the inclined scale.

**Figure 2:**
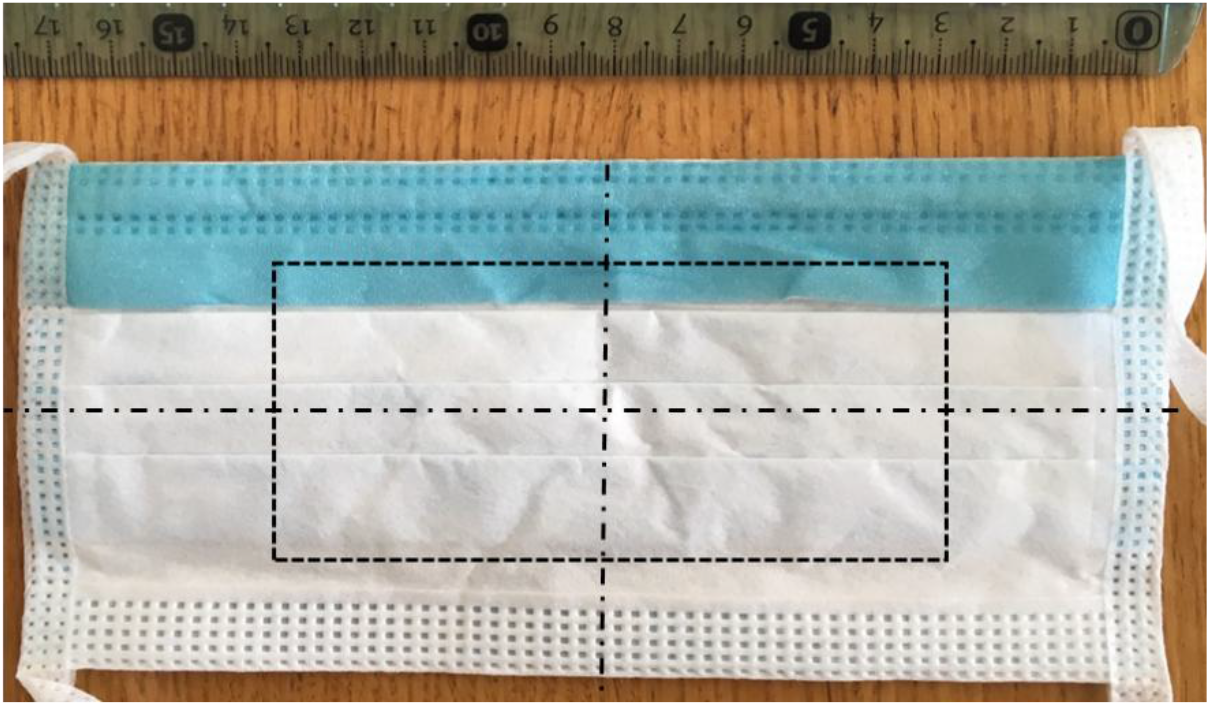
Indication of the test control surface area for the masks, width 110 mm and height 55 mm.

#### Air flow rate measurement

An Alcock/Ricardo Viscous Flow Air Meter (type 50 cfm VFAM) was used to measure the volumetric flow rate through the test rig. The meter uses a resistive laminar flow element to generate a pressure drop across the meter that has a strong linear relationship with volumetric flow rate for a given meter calibration factor, *k (*m^3^s^−1^Pa^−1^). A temperature correction for viscosity effects is applied using Sutherland’s law to evaluate the volume flow rate, *Q_Air_* (m^3^s^−1^) as follows:

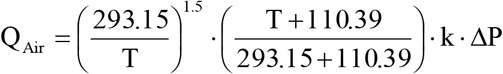

where 293.15 K is the reference temperature, 110.39 is Sutherland’s constant, T (K) is the VFAM temperature, and ΔP (Pa) is the VFAM pressure reading. It is not generally necessary to correct for ambient humidity within the flow rate ranges used. The meter is capable of measurements up to 25 ls^−1^ with an accuracy of +/− 0.05 ls^−1^. The pressure drop across the meter was measured using a KIMO type CP304 piezoresistive, differential pressure transducer (+/− 0.5 % reading or +/− 10 Pa). Temperature at the VFAM was measured using a type K thermocouple (+/− 1 °C). Under pandemic lockdown conditions it was not possible to re-calibrate the flow meter prior to testing. Instead, the linearity of the device was evaluated by comparing the previous calibration data (supplied by Young Calibration Limited, UK) against data obtained at the same flow rates, alternately using the differential pressure transducer and an inclined liquid manometer to record the pressure. The results confirmed that both pressures were equal. This concluded that the volumetric flow rates were consistent with the calibrated values, given that the VFAM element was clean and the device does not have any moving or electronic components subject to drift. A linear extrapolation of the calibration data, based upon the assumptions that the calibration curve passes through the origin and that the flow remains laminar throughout the range of tests, was used to determine the pressure drop required to ensure the correct flow breathing flow rates for testing. Repeatability in the test results also confirmed the reliability of the test equipment for comparative purposes.

#### Aerosol generation

A pharmaceutical nebuliser (Medix Econoneb) was used to generate a fine mist of tap water with a dynamic flow rate of between 7 lmin^−1^ and 9 lmin^−1^ at 138 kPa. The particle MMAD is 3.4 μm and 72% of particles are < 5 μm (measured in accordance with BSEN 13544–1). The nebuliser chamber was inserted into the base of the flow chamber where the mist formed a homogenous mixture with the incoming air.

### Experimental Procedure

**Part 1**. Testing of the masks was carried out at two mean, inspiratory, steady-state, flow (MIF) rates of 30 lmin^−1^ and 95 lmin^−1^ (see EN 149–2001+A1:2009; 2009). These were adopted in order to evaluate the masks at conditions representative of difficult working conditions; deep breathing in a clinical setting and vigorous effort, such as in an emergency response setting respectively (Grinshpun et al. 2009). The airflow direction was from the outside of the mask to the inside, mimicking inhalation. All the masks were clamped flat with all 3 layers utilised. The control area was selected to incorporate the main construction elements of the masks (so as to incorporate all variations in the quality) in a similar manner to the EN 14683: 2019 (2019) differential pressure test (and also to cover the mouth and nose areas of the wearer). This representative area (cut into the flow plates) measured 60.5 cm^2^ (see Figure 2) for all of the masks.

An initial phase of testing was conducted with the CEN EN approved Control masks to establish a suitable methodology, evaluate robustness and repeatability of the measurements. Sixteen tests at the two flow rates were conducted using four masks. The same tests were repeated at the end of testing of all the sample masks and showed the results were reproducible. A fixed mass flow rate approach, opposed to a fixed pressure drop method, was adopted. The following simple methodology was devised:

1. A mask was attached to the mounting plate and clamped in position in the flow chamber.
2. All instruments were zeroed.
3. Motive compressed air was regulated to 4 bar and the eductor orifice varied until the required pressure drop was noted on the KIMO differential pressure display at the VFAM (indicating the desired volume flow rate).
4. The display was left to stablise over a period of approximately 30 seconds. (The test rig instruments proved steady and repeatable).
5. The pressure drop across the masks was then read off the scale on the inclined manometer.
6. The higher flow rate was selected using the eductor and the measurements repeated.
7. The lower and higher flow rates were then selected again, and the results recorded once more.
8. The mask was removed and replaced with the next mask from the same sample until all samples were tested.

At least 4 repeat tests were carried out from each sample of masks selected at random. The laboratory conditions were at room temperature (24 °C recorded at the VFAM for all tests) and 1015 mbar ambient pressure. In excess of 60 tests in total were carried out for the inhalation resistance tests. The results were recorded as volumetric flow rate against pressure drop and averaged over the 4 samples within each batch. Whereas the absolute values of pressure drop are unlikely to correlate with other testing approaches, the observed, indicative trends and relative performance of the masks is key to this approach.

**Part 2**. In a further test, designed to evaluate the suitability of the rig for future experiments, the clogging resistance of the masks to small water droplet clouds was evaluated for the CEN EN approved Type IIR fluid resistant surgical masks. The experimental setup was carried over from previous tests for a flow rate case of 95 lmin^−1^. Water droplets from a pharmaceutical nebulizer (specification outlined above) were fed continuously into the flow chamber, homogenously mixed upstream of the mask. This was visually confirmed by video recording through the back of the spray chamber. The flow rate was maintained constant and the pressure drop was recorded every 5 minutes up to a maximum time just short of 30 minutes.

### Preliminary Results

**Part 1**. The mean pressure drop for all the tests for each group of masks was calculated for each flow rate condition. The pandemic conditions limited the time available to conduct the tests and therefore the sample size. However, the standard deviation and coefficient of variation (CoV) in the datasets were evaluated as an indicative result of test repeatability. For the pressure drop measurements, the standard deviation varied between 1 to 6.5 Pa for the ‘deep breathing’ conditions and 6 to 21 Pa for the ‘vigorous effort’ conditions, The CoV values ranged between approximately 5 % to 12% for the low flow rate case and between 6 % and 12 % for the higher flow rate. These results are to be expected given the experimental uncertainty in setting the required flow rate at the lower rate could be of the order of 10%. Samples A and B consistently showed the highest values at both flow rates. Sample C exhibited the lowest CoV combined over both flow rates. All the samples were then ranked against the CEN EN certified FRSM Type IIR mask in terms of their inhalation resistance as estimated by pressure drop. The results for the mean pressure drop values, at the two different flow rates, for all the samples tested, are presented in Figure 3 below with +/− 1 standard deviation in pressure drop included for reference. The results indicate that none of the sample masks matched the pressure drop of the Type IIR mask at either condition; a mean pressure drop of approximately 255 Pa for the upper case, compared to a range of approximately 55 Pa to 140 Pa for the other samples. For the ‘deep breathing’ conditions, masks A to D presented mean pressure drop values of approximately 19 Pa to 42 Pa whereas the Type IIR mask had a mean pressure drop value of approximately 72 Pa.

**Figure 3:**
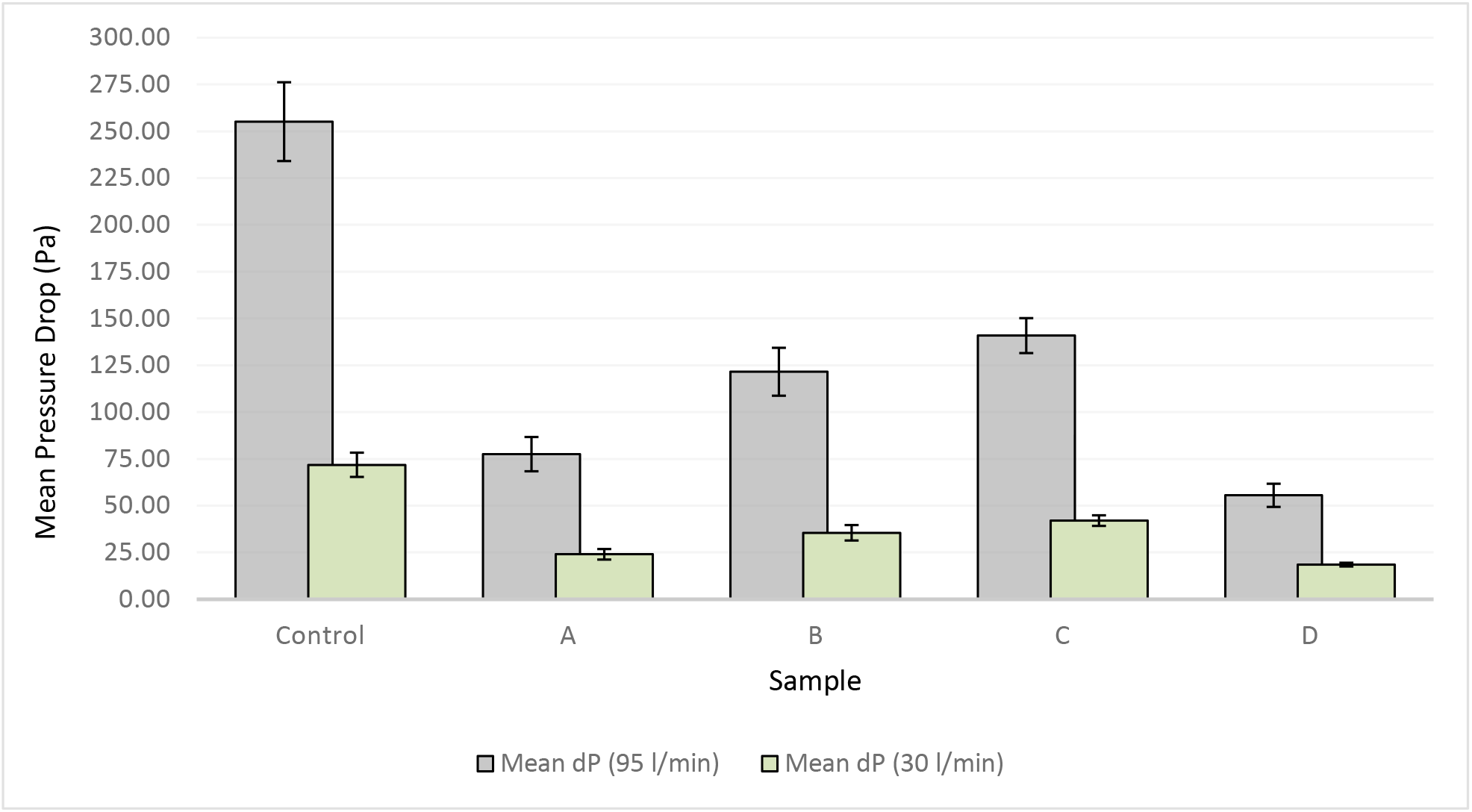
Comparison of the measured mean pressure drop between the sample and control masks.

All the samples were then ranked against the CEN EN certified FRSM Type IIR mask in terms of their inhalation resistance as estimated by pressure drop. The results as a percentage normalised to the maximum pressure drop observed for the certified control set are shown in Tables 3a, b.

**Table 3a:**
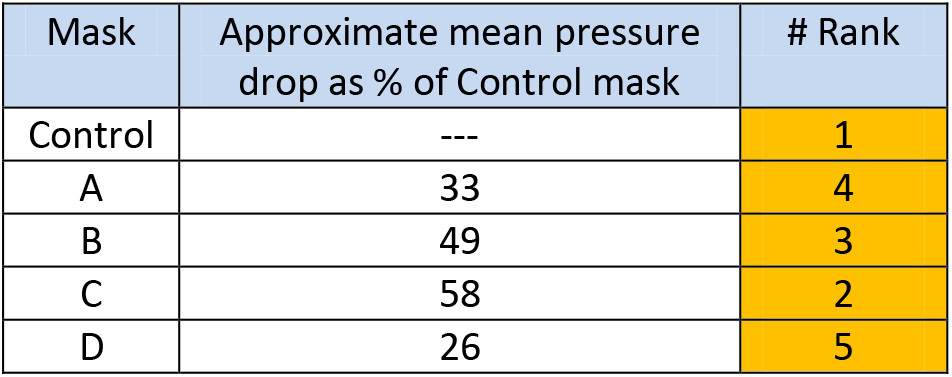
Ranking of the masks compared to control mask at 30 lmin^−1^.

| Mask | Approximate mean pressure drop as % of Control mask | # Rank |
| --- | --- | --- |
| Control | --- | 1 |
| A | 33 | 4 |
| B | 49 | 3 |
| C | 58 | 2 |
| D | 26 | 5 |

**Table 3b:**
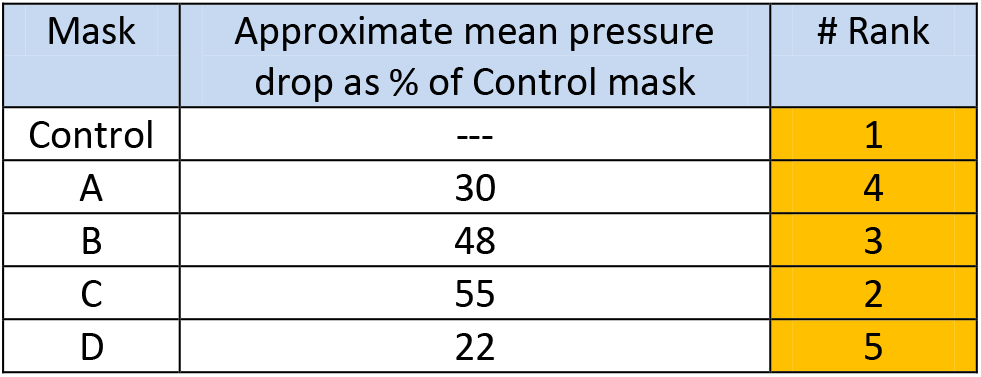
Ranking of the masks compared to the control mask at 95 lmin^−1^.

From Figure 3 and Tables 3a, b, it is clear that the control Type IIR mask consistently outperformed the other masks in terms of pressure resistance to flow for both conditions. Samples from each batch of masks performed consistently at each of the test conditions; mask A ranked at 33% (at 30 lmin^−1^) and 31% (at 95 lmin^−1^) of the values of the control mask; mask B ranked at 49% (at 30 lmin^−1^) and 48% (at 95 lmin^−1^) of the values of the control mask; mask C, likewise, 58% (at 30 lmin^−1^) and 55% (at 95 lmin^−1^) of the values of the control mask; mask D ranked lowest at 26% (at 30 lmin^−1^) and 22% (at 95 lmin^−1^) of the values of the control mask. Therefore, the consistent ranking for both high and low flow rates indicated good robustness in the experimental approach. It is interesting to note that one mask in the control sample was used up to 8 times in a series of separate tests (including during setup tests) and started to demonstrate a consistently lower pressure drop of around 10% compared with its first use. It was noted that this could be due to the clamping force placed upon the mask eventually distorting the pleats, thereby exposing more surface area.

**Part 2**. The results of the rig evaluation tests for clogging, using the nebulized water droplet cloud, are presented in Figure 4 below. The estimated experimental uncertainty in the pressure measurement using the inclined manometer is included as an error bar. The results show a very small increase in pressure after approximately 15 minutes of continuous operation, but care must be taken in interpreting such results for several reasons. Firstly, the measurements are very small, and the uncertainty is likely of the order observed in the main experiments. Secondly, the FRSM mask is a splash resistant mask that is not designed to filter particles below approximately 3 µm. In this case, more than 70% of the droplets generated by the nebuliser are less than 5 µm in diameter and a large majority are likely to pass straight through. Nonetheless, it is useful to include these results here as the experiment has shown that there is potential to pursue a simple test such as this with, for example, FFP/N95 type respirators (see Table 1), dyes and imaging.

**Figure 4:**
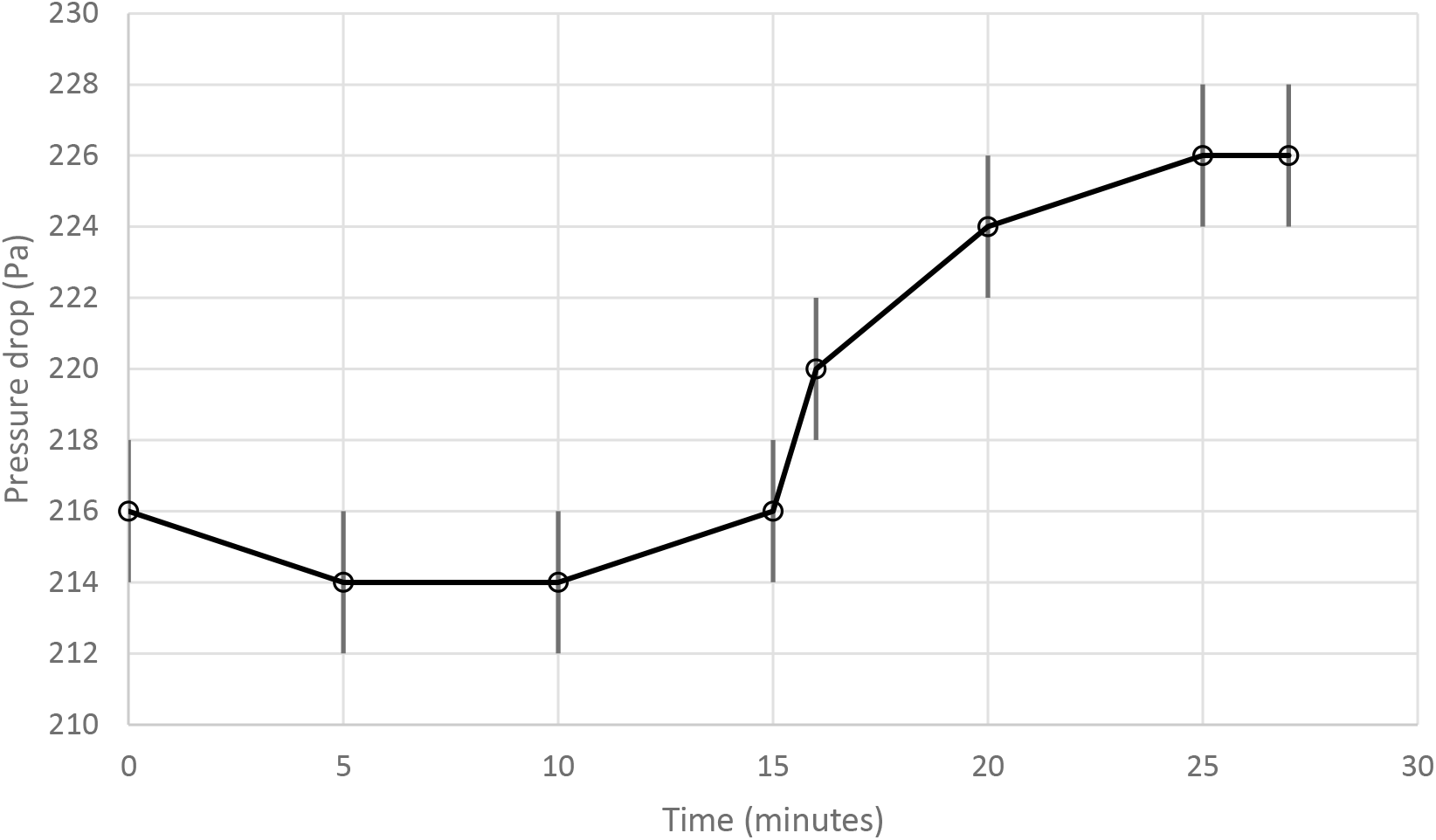
Evolution of pressure drop with time for CEN EN approved FRSM Type IIR masks for a fixed flow rate of 95 lmin^−1^ for the case of continuous upstream water injection of droplets of MMAD 3.4 µm.

## Evaluation, Impact and Scalability

All the sample masks performed well below the standard of the CEN EN control Type IIR mask for all conditions tested. However, masks B (48–49%) and C (55–58%) performed far better in these tests than masks A (30–33%) and D (22–26%). Masks A to D, based on their inhalation resistance performance, would not be recommended for use in a surgical setting. (Anecdotally, masks A and D performed less effectively than a home-made mask). However, masks B and C may potentially be utilised in more appropriate non-medical settings when professionally assessed. In a low resource, public health emergency or indeed in a low resource environment, being able to make an informed decision without undergoing the certification process saves time and expense. It must be noted that this test protocol does not, and is not intended in any way, to replace the certification process for masks. However, with the likely increase in the adoption of masks, mandatory or otherwise and the potential for a high proportion to fall short of certification standards, an approach such as adopted here may prove useful in gauging the potential usability of these masks. A potential decision flow path as illustrated in Figure 5 below could be envisaged.

**Figure 5:**
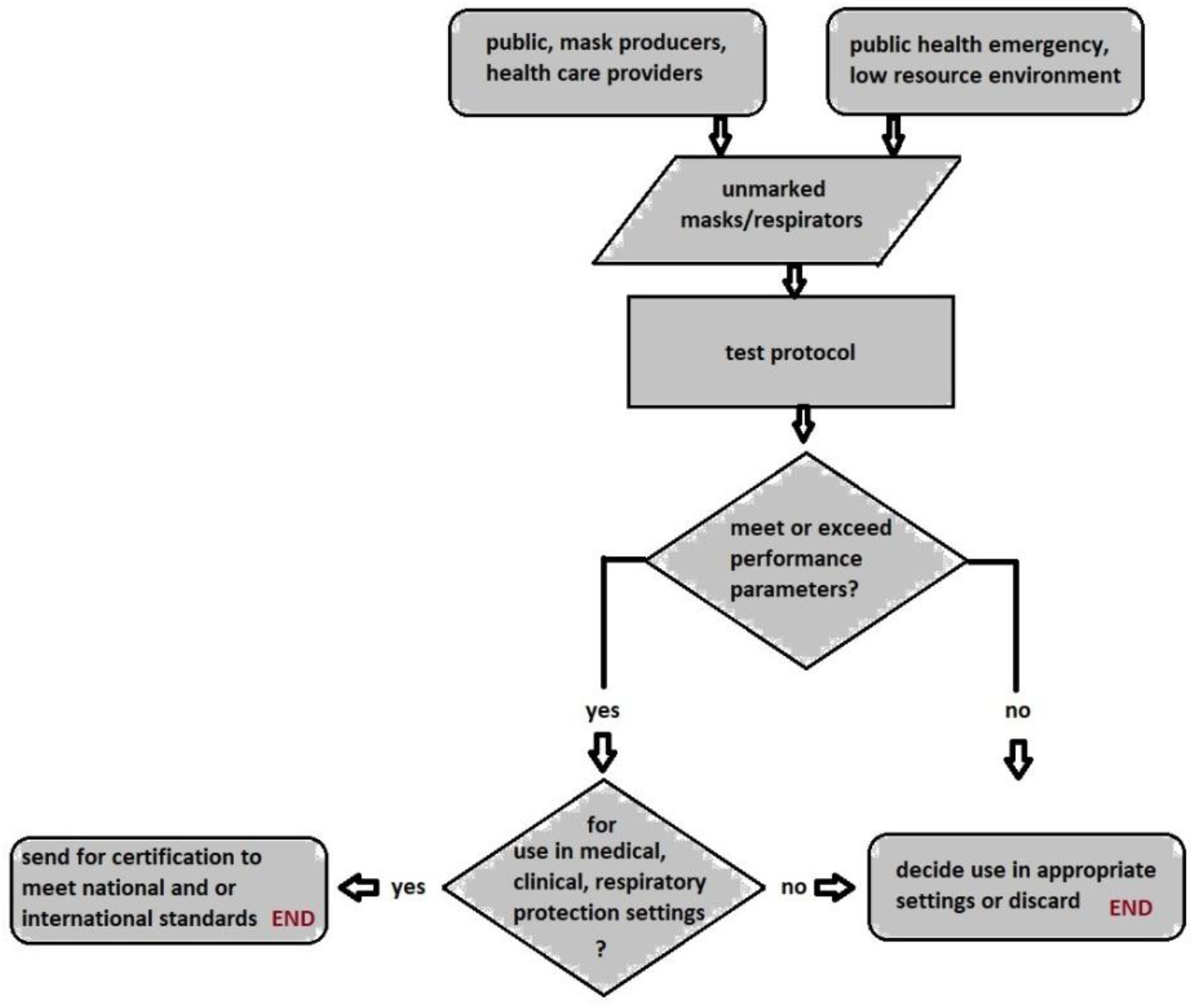
A possible decision path for using the rapid assessment protocol.

## Conclusions and further work

A rapid assessment method for medical face masks, based upon their steady-state inhalation resistance to air flow, has been developed and experimentally tested. The method works by drawing air through the masks and a series of pipes and flow devices at a known flow rate condition representative of human activities; deep breathing and vigorous effort. The resistance to the air flow through the device is measured as the pressure drop across a specified portion of the mask. This approach mimics air inhalation through the mask. A sample of certified Type IIR FRSM surgical masks were used as the control reference for the evaluation of the performance of 4 different, randomly selected batches of uncertified but physically similar types of medical masks. The results showed that all the masks tested produced pressure drop values in the range of 22% to 58% of the control set and that their performance was consistently observed at all test conditions. Within the group of masks tested, two sets (between 48% and 58% of the reference set) showed the potential to be professionally assessed for appropriate deployment in a suitable setting. The experimental setup, including component design and test rig evaluation was carried out in several days. In excess of 60 measurements were carried out within 8 hours. The methodology provides a rapid, low resource air inhalation test that can be used to assess the best route forward for masks early in the allocation stage; send for certification; deploy in medical or other more appropriate settings. Such an approach has very recently been proven useful during the COVID-19 pandemic public health emergency in the UK but would be suitable in low resource regions worldwide. The test does not replace any National or International certification standards for masks.

Further work is focused upon the development of the test rig, instrumentation and experimental method with the view to providing a benchmark for a smaller testing device that can be deployed in the field. Overcoming some of the observed limitations, as mentioned previously, such as flat clamp testing, rapid mask exchanging and introducing transient (pulsing) flow rates will be reviewed. In addition, further development of the method for clogging, as discussed briefly above, extended to polydisperse droplet particle size distributions, is also currently being considered.

## Data Availability

All the data have been reported

## Acknowledgements

Many people helped us to carry out this project, without a budget, from conception to completion in 4 days. Mr Barney Harle and Mr Simon Gardiner of Manchester City Council donated the masks, provided advice, certification protocols and reviewed the results. Professor Hector Iacovides and Mr Dennis Cooper (University of Manchester) reviewed the test protocol and results. Ms Emilian Kayone (BEng final year student at the University of Brighton) who remotely provided information about the test rig dimensions. Professor Debra Humphris, Vice-Chancellor of the University of Brighton and Professor Taraneh (Tara) Dean, Pro-Vice-Chancellor (Research and Enterprise) and Dr John Taylor, Head of School of Computing, Engineering and Mathematics, for granting special permission to use the laboratories. The University Estates and Security Teams, for opening up the facilities and organising electrical power, compressed air, water and other utilities in the Advanced Engineering Building at the University of Brighton.

## Author contributions

Investigation: NE, OE. Methodology: NE, SB, NM. Project Administration: NE, OE, SB. Writing – draft: NE, SB. Writing – review: NE, SB. Conceived and designed the experiments: SB, NM, BW. Performed the experiments: SB, BW. Analysed the data: SB, NM, NE. Designed and constructed the flow chamber: NM, SB.

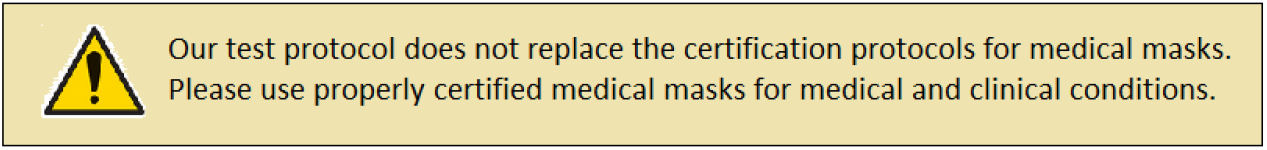

